# Development and Cross-Validation of a Short Questionnaire to Evaluate Self-Reported Positive Health; A Cross Sectional Panel Study of Structural Validity Among a General Dutch Population

**DOI:** 10.1101/2024.08.26.24312579

**Authors:** Lenny M.W. Nahar-van Venrooij, Margot J. Metz, Marja van Vliet, Vera P. van Druten, Babette C. van der Zwaard

## Abstract

**Objectives:** In this study it was aimed to further develop and cross-validate a short questionnaire to measure self-reported Positive Health in general (Dutch) populations for evaluative purposes, stemming from the original 42 items of the My Positive Health dialogue tool (MPH). Positive Health refers to ‘health from the perspective of patients and citizens’ following the concept of Huber et. al. **Design and setting**: A cross sectional study was performed among a panel representative for the general adult Dutch population living at home.

**Participants:** Response rate was 76%, 1327 of a total of 2457 respondents were female, and mean age (year) was 53.3 ± 17.8.

**Methods:** First, item reduction was carried out through content discussions following statistical output retrieved from factor structures and loadings, inter-item correlations (IIC) and internal consistency (Cronbach’s alphas). Next, among the other half of the study population, measurement properties for the developed short questionnaire were calculated using goodness of fit indices from confirmatory factor analyses (CFA).

**Results:** The item reduction process (n=1199) resulted in a questionnaire of 22 items (PH22) with a four-factor structure and explained variance of 62.4%. Cronbach’s alphas were 0.84, 0.92, 0.81, and 0.78 for the renamed factors ‘Physical fitness’ (5 items), ‘Contentment with life’ (9 items), ‘Daily life management’ (5 items) and ‘Future perspective’ (3 items), respectively. Cross validation (n=1258) showed adequate goodness of fit indices of the PH22, based on both first– and second-order CFA. The scores of the PH22 were normally distributed. No floor or ceiling effects were present.

**Conclusions:** A short 22 item questionnaire to measure self-reported Positive Health in a general (Dutch) population for evaluative purposes such as scientific or policy research at Positive Health or patient-centered interventions was developed and cross-validated, named PH22. This study supports its structural validity. To use this questionnaire in practice its test-retest reliability and responsiveness should be known also. Future research has to reveal this.

*Strengths and limitations of this study:*

- The main strength of this study was that the choice to keep or remove an item during the development of the short Positive Health questionnaire was not only based on statistical output such as factor loadings, but combined with thorough content discussion by the expert team and judgement of inter-item correlations and internal consistency.
- This study is robust in terms of its large sample size, the high response rate and the representativeness of the general Dutch population.
- Development of the short Positive Health questionnaire was based on the items of the My Positive Health dialogue tool, which is widely used in the Netherlands.
- It can be argued that content discussion is less objective or transparent to follow than statistical output. To overcome this, the results from the content discussion were thematized and each step of the item reduction process thoroughly reported.
- Choices made by the expert team, might have been more support-based if more representatives were included in the content discussion, i.e., if focus groups were organized. Nevertheless, the members of the research team represent different backgrounds and relevant expertise. Moreover, it should be realized that the basic set of items of the My Positive Health dialogue tool was based on health indicators retrieved from a large study among various stakeholders and judged relevant.

## INTRODUCTION

Since the concept of Positive Health was introduced in the Netherlands, a mind shift unrolled among healthcare workers and beyond. The approach of health as a state of complete physical, mental and social well-being as formulated in the constitution of the World Health Organization(1) changed to a more dynamic approach of health focusing on self-management and the ability to adapt to physical, mental and social challenges during life(2). This new vision on health is being integrated among all kinds of domains and political agendas within the Netherlands and abroad(3).

To support the applicability of this vision on health in daily healthcare practice, the dialogue tool My Positive Health (MPH)(4) was developed. The content of this dialogue tool was derived from a large mixed methods study with interviews into the perceptions about health among different stakeholder groups such as patients, citizens, and healthcare professionals(5). This inductive, bottom-up approach enabled the researchers to gain a thorough insight into the perceptions about health. From these perceptions 32 aspects emerged, representing indicators for (positive) health(5). Accordingly, these aspects were thematized among six dimensions named: bodily functions, mental functions and perception, spiritual existential dimension, quality of life, social and societal participation and daily functioning. This operationalization of health was called *Positive Health*, and from here the 42-item MPH dialogue tool was developed. This MPH tool aims to support the conversation about Positive Health between patient and care worker and stimulate self-reflection(4).

At an individual, organizational, community, regional and national level, the concept (broad and dynamic vision on health) and method (MPH tool and dialogue) are increasingly integrated. The Dutch government considers Positive Health a promising approach to promoting well-being and handle the increasing burden of disease(6). To assess the effectiveness of working with this Positive Health approach, the need for an instrument to measure self-reported Positive Health has been arising(7,8). Although the MPH is a relevant dialogue tool for the conversation about health(3), it should be emphasized that the MPH is not obviously useful for measuring purposes; the item grouping among the six dimensions of the MPH tool was not the result of a study aiming to assess structural validity in order to develop an outcome measure instrument.

To our knowledge, two instruments were developed for this measuring purpose; the Positive Health measurement scale with 17 items (PH17)(9,10) and the Positive Health measurement tool using all 42 dialogue items (PH42)(11). These two instruments face some limitations. Although measurement properties for the PH17 seemed adequate(10).the initial item selection of the PH17 took place among citizens in just one part of the Netherlands and response rate was low (25%)(9), questioning the generalizability of their results. Even more important, the methodological approach for item reduction included judgement of factor loadings, but without, simultaneously, content discussion and judgement of inter-item correlations and maintaining acceptable internal consistencies as recommended by others(12). Without these steps relevant items might be deleted, and shortchange its content and discriminant validity. The other instrument, the PH42, was developed among a representative general population(11), but consists of 42 items which might not be preferable for all practices. From practical and methodological perspectives, it is preferable to use a shorter questionnaire, which requires less effort and results in higher response rates, especially important during repeated measurements needed to evaluate (positive) health or patient-centred interventions.

The aim of this study was to develop and cross-validate a short questionnaire to measure self-reported Positive Health for evaluation purposes in scientific or policy research at Positive Health promoting, or patient-centered interventions in general populations. Its structural validity was assessed and the more extensive method for item reduction was applied among a representative study population. The conditions set were that the questionnaire had to contain the original items of the MPH dialogue tool to retain its recognizability with daily practice and with Positive Health as operationalised by Huber et. al.(5), referring to ‘health from the perspective of patients and citizens’.

## METHODS

### Study design and participants

In this paper, we make use of data from the LISS panel (Longitudinal Internet studies for the Social Sciences) managed by the non-profit research institute Centerdata (Tilburg University, the Netherlands). The LISS panel consists of a representative sample of approximately 7,000 individuals from 5,000 households from the general Dutch population. The panel is based on a true probability sample of households drawn from the population register by Statistics Netherlands(13). LISS panel members complete monthly online questionnaires and are paid for each completed questionnaire. To become a LISS panel member, at least one person in the household has to be proficient in the Dutch language. To minimize selection bias, households were provided with a computer and internet connection if they could otherwise not participate. Response rates for this panel are high (>80%). More information about the LISS panel can be found at: www.lissdata.nl.

To answer our research question a cross sectional study was performed among a random selection of members from the LISS panel. From this panel, 2,500 adults (≥18 years), one per household, were randomly selected to participate. The process of item reduction and cross-validation were carried out in two randomly split samples of this study population. Ethical review was conducted by the METC Brabant (Tilburg, the Netherlands, study number NW2024-15).

This study was reported according to the COSMIN Reporting Guideline(14) recommended for studies that evaluate the measurement properties of patient-reported outcome measures (PROMs). The terms dimension and factor are used interchangeably.

### Data collection and administration

During November 2020 the selected study population was asked to complete the original 42 items of the My Positive Health questionnaire (MPH) (see Additional file A) receiving one reminder after 2 weeks. The same as the original MPH dialogue tool the items were introduced per dimension using the original introduction, answer options and icons of the dialogue tool(4). In contrast to the original tool the respondents did not see their results among a spiderweb. Respondents completed the electronic questionnaire at home using the regular internet platform of LISS receiving a private link. Characteristics of the study population such as gender, age, level of education and health care use were available from the regular LISS panel HEALTH survey (https://www.lissdata.nl/research/liss-core-study).

### My Positive Health (MPH) dialogue tool

The MPH consists of 42 statements about Positive Health, representing the 32 indicators for (positive) health as assessed by Huber et al.(5). For practical use, they were formulated to a simple language level (B1). The statements are scored on an 11-point Likert scale ranging from 0 ‘completely disagree’ to 10 ‘completely agree’. Higher scores indicate better health. Also, the six dimensions (bodily functions, mental functions and perception, spiritual existential dimension, quality of life, social and societal participation and daily functioning) are visualised in a spider web with six axes, representing the dimensions and ranging from value 0 (in the centre for poor) to 10 (on the periphery, for excellent). The self-reported MPH questionnaire takes 10-20 minutes to complete. Over the last years it was shown by various users (citizens, patients and professionals) that the MPH was a relevant dialogue tool including comprehensiveness and comprehensibility of the items, response options, and instructions(3).

### Preconditions for the short Positive Health questionnaire to be developed

Preconditions formulated by the research team for an useful self-reported questionnaire to measure Positive Health were; 1) a multidimensional structure was held to ensure a broad representation of health conform literature(5,15), 2) items were not reformulated to keep recognizability with the specific Positive Health dialogue approach according to MPH(4), 3) to hold model stability each dimension contained at least three items (12), and 4) the short questionnaire contained a maximum of about 20 items to be user-friendly.

### Statistical analyses

#### Development: Process of item reduction

Prior to this study, Van Druten et al. developed the measurement tool PH42 (11). They assessed the factor structure of the 42 original items of the dialogue tool MPH. This resulted in a model with a six-factor structure including all 42 items with an explained variance of 68%, no inter-items correlations > 0.9, factor loadings ranging from 0.36 to 0.94, Cronbach’s alpha’s ranging from 0.74 up to 0.97, and acceptable fit indices. This study of Van Druten et al. was based on the same dataset as our study. Their results (see Additional file B-C) were the starting point for the item reduction process of our study. We used the same settings to assess dimensionality during the process of item reduction: extraction method; Principal Component Analysis (PCA)(16–18), rotation method; Oblimin with Kaiser Normalisation, and eigen value >1.0 using SPSS V27.0. Analyses were performed on similar randomly split half of the study population (n=1199).

The following steps of the item reduction process were taken conform the methodology published by De Vet and Terwee ^12,15^. Content discussions initiated through statistical output were performed in different rounds with experts taking part in the research team. First, the items of the PH42 were assessed per factor on low (<0.2; i.e. possibly unrelated to the construct) and high (>0.7; i.e. possibly overlapping and thus redundant in the construct) inter-item correlations(12). Based on content discussion low or highly correlated items were held or removed. Then, PCA was performed. Items that hardly loaded at all on any of the factors were considered for deletion. A minimum factor loading of 0.5 was taken as threshold(12). Also, items loading >0.32(12,19) on more than one factor were discussed. Based on content discussion, items were held or removed. Content was leading, meaning that for some items, high correlations or low factor loadings might be accepted. Items were deleted one by one repeating PCA every step, because deletion of one item might change structures or loadings of other items(12). Final decisions to delete an item were combined with judgement of consequences for internal consistencies (Cronbach’s alpha) aimed between 0.7-0.9 (12).

#### Cross validation

To assess goodness of fit of the developed short Positive Health questionnaire, confirmatory factor analyses (CFA) was performed in the second half of the study population (n=1258) CFA for normal continuous data with maximum likelihood (ML) as estimation method was used (R Lavaan 0.6.14)(20). Goodness of fit indices included; chi-square (X^2^) (a non-significant X^2^ is desirable, however in a large sample, the X^2^ is usually significant), comparative fit index (CFI), root mean square error of approximation (RMSEA) and standardized root mean square residual (SRMR). Indicators of model fit were(12,21); CFI values between 0.90 and 0.95 with >0.95 indicating superior model fit, RMSEA values <0.05 represent good fit, 0.05-0.08 acceptable fit, >0.08 medium fit and >0.1 poor fit, and SRMR value of <0.08 representing good fit. To assess if the item scores of the questionnaire fit the factor sum scores first-order CFA was executed. To investigate if the factor sum scores fit the total sum score of the questionnaire as well, second-order CFA was executed(12,20).

#### Scores of the developed questionnaire

Last, the distribution of the total and factor sum scores of the developed questionnaire were described; mean, median, standard deviation, minimum, maximum, skewness and kurtosis (< –1 and > 1), and floor and ceiling effects (≥ 15% of the respondents scores lowest or highest possible scores, respectively(22)).

### Sample size calculation

Size of both randomly split subgroups (n=1199, n=1259)(11) was adequate to apply PCA and CFA; rule of thumb is that four to ten respondents per item of the questionnaire are included, with a minimum of 100 (23).

### Patient and public involvement

Patients or the public were not involved in the design, or conduct, or reporting, or dissemination plans of our research.

## RESULTS

### Participants

The response rate was 76% with 777 respondents not responding. Twelve respondents not completing the questionnaire completely were excluded, leaving 2457 respondents for the analyses; 54% female, mean age (years) 53.3 ± 17.8, 39.9% high level of education, and 39.8% visited a medical specialist at the hospital, psychiatrist, psychologist or psychotherapist last 12 months. Next, the study population was randomly split: n=1199 and n=1258, in which the process of item reduction and cross-validation was carried out, respectively.

### Development: Process of item reduction

LNvV, BvdZ, MM and MvV participated at six research meetings of an hour between May and August 2023 concerning the item reduction process; content discussion and interpretation of the statistical output. During **round 1** interitem correlations were explored for the six-factor structure of the PH42 (see Additional file C;. From all factors four contained half or more items that were too highly (>0.7) correlated to another item: Factor 1 (11 out of 13), factor 2 (4/8), factor 3 (2/7), factor 4 (5/8), factor 5 (0/3) and factor 6 (2/3), respectively. Two of all items correlated low (<0.2) with each other but adequately with the other items; factor 2 (2/8). First, the items with interitem correlations >0.8 were discussed on their content, next those items with correlations >0.7. Initiated by these high correlations content discussion led to choices for deletion of an item for various reasons such as inadequate formulation of the statement, not being inclusive or (not) being specific. In Table 1 detailed information about the choices made per item are shown.

**Table 1.**
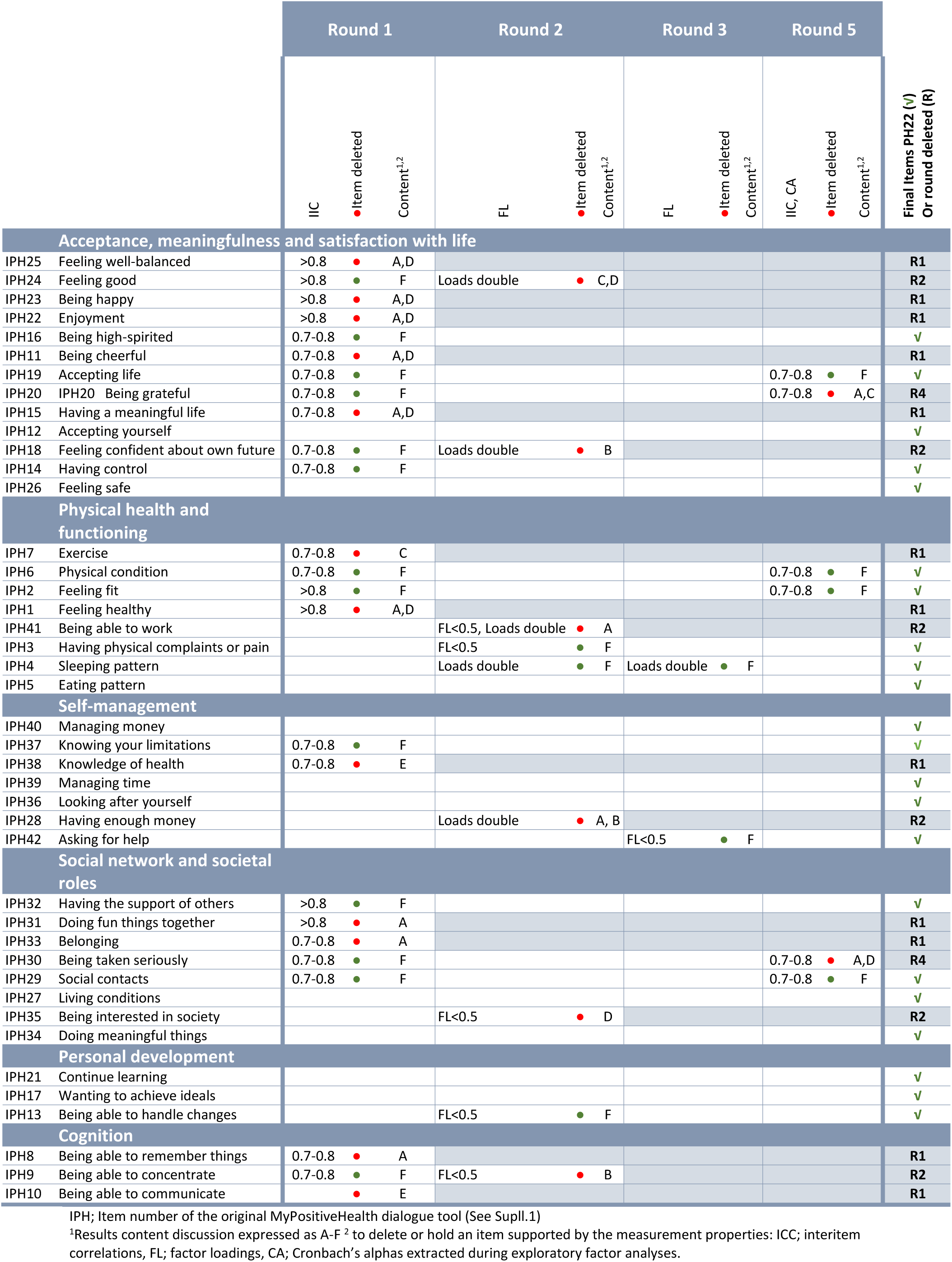
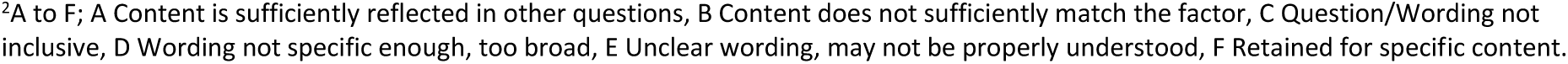
Process of item reduction with the PH42 questionnaire as starting point.

|  |  | Round 1 |  |  | Round 2 |  |  | Round 3 |  |  | Round 5 |  |  | Final Items PH22 (✓)<br>Or round deleted (R) |
| --- | --- | --- | --- | --- | --- | --- | --- | --- | --- | --- | --- | --- | --- | --- |
|  |  | IIC | Item deleted | Content<sup>1,2</sup> | FL | Item deleted | Content<sup>1,2</sup> | FL | Item deleted | Content<sup>1,2</sup> | IIC, CA | Item deleted | Content<sup>1,2</sup> |  |
| Acceptance, meaningfulness and satisfaction with life |  |  |  |  |  |  |  |  |  |  |  |  |  |  |
| IPH25 | Feeling well-balanced | >0.8 | ● | A,D |  |  |  |  |  |  |  |  |  | R1 |
| IPH24 | Feeling good | >0.8 | ● | F | Loads double | ● | C,D |  |  |  |  |  |  | R2 |
| IPH23 | Being happy | >0.8 | ● | A,D |  |  |  |  |  |  |  |  |  | R1 |
| IPH22 | Enjoyment | >0.8 | ● | A,D |  |  |  |  |  |  |  |  |  | R1 |
| IPH16 | Being high-spirited | 0.7-0.8 | ● | F |  |  |  |  |  |  |  |  |  | ✓ |
| IPH11 | Being cheerful | 0.7-0.8 | ● | A,D |  |  |  |  |  |  |  |  |  | R1 |
| IPH19 | Accepting life | 0.7-0.8 | ● | F |  |  |  |  |  |  | 0.7-0.8 | ● | F | ✓ |
| IPH20 | IPH20 Being grateful | 0.7-0.8 | ● | F |  |  |  |  |  |  | 0.7-0.8 | ● | A,C | R4 |
| IPH15 | Having a meaningful life | 0.7-0.8 | ● | A,D |  |  |  |  |  |  |  |  |  | R1 |
| IPH12 | Accepting yourself |  |  |  |  |  |  |  |  |  |  |  |  | ✓ |
| IPH18 | Feeling confident about own future | 0.7-0.8 | ● | F | Loads double | ● | B |  |  |  |  |  |  | R2 |
| IPH14 | Having control | 0.7-0.8 | ● | F |  |  |  |  |  |  |  |  |  | ✓ |
| IPH26 | Feeling safe |  |  |  |  |  |  |  |  |  |  |  |  | ✓ |
| Physical health and functioning |  |  |  |  |  |  |  |  |  |  |  |  |  |  |
| IPH7 | Exercise | 0.7-0.8 | ● | C |  |  |  |  |  |  |  |  |  | R1 |
| IPH6 | Physical condition | 0.7-0.8 | ● | F |  |  |  |  |  |  | 0.7-0.8 | ● | F | ✓ |
| IPH2 | Feeling fit | >0.8 | ● | F |  |  |  |  |  |  | 0.7-0.8 | ● | F | ✓ |
| IPH1 | Feeling healthy | >0.8 | ● | A,D |  |  |  |  |  |  |  |  |  | R1 |
| IPH41 | Being able to work |  |  |  | FL<0.5, Loads double | ● | A |  |  |  |  |  |  | R2 |
| IPH3 | Having physical complaints or pain |  |  |  | FL<0.5 | ● | F |  |  |  |  |  |  | ✓ |
| IPH4 | Sleeping pattern |  |  |  | Loads double | ● | F | Loads double | ● | F |  |  |  | ✓ |
| IPH5 | Eating pattern |  |  |  |  |  |  |  |  |  |  |  |  | ✓ |
| Self-management |  |  |  |  |  |  |  |  |  |  |  |  |  |  |
| IPH40 | Managing money |  |  |  |  |  |  |  |  |  |  |  |  | ✓ |
| IPH37 | Knowing your limitations | 0.7-0.8 | ● | F |  |  |  |  |  |  |  |  |  | ✓ |
| IPH38 | Knowledge of health | 0.7-0.8 | ● | E |  |  |  |  |  |  |  |  |  | R1 |
| IPH39 | Managing time |  |  |  |  |  |  |  |  |  |  |  |  | ✓ |
| IPH36 | Looking after yourself |  |  |  |  |  |  |  |  |  |  |  |  | ✓ |
| IPH28 | Having enough money |  |  |  | Loads double | ● | A, B |  |  |  |  |  |  | R2 |
| IPH42 | Asking for help |  |  |  |  |  |  | FL<0.5 | ● | F |  |  |  | ✓ |
| Social network and societal roles |  |  |  |  |  |  |  |  |  |  |  |  |  |  |
| IPH32 | Having the support of others | >0.8 | ● | F |  |  |  |  |  |  |  |  |  | ✓ |
| IPH31 | Doing fun things together | >0.8 | ● | A |  |  |  |  |  |  |  |  |  | R1 |
| IPH33 | Belonging | 0.7-0.8 | ● | A |  |  |  |  |  |  |  |  |  | R1 |
| IPH30 | Being taken seriously | 0.7-0.8 | ● | F |  |  |  |  |  |  | 0.7-0.8 | ● | A,D | R4 |
| IPH29 | Social contacts | 0.7-0.8 | ● | F |  |  |  |  |  |  | 0.7-0.8 | ● | F | ✓ |
| IPH27 | Living conditions |  |  |  |  |  |  |  |  |  |  |  |  | ✓ |
| IPH35 | Being interested in society |  |  |  | FL<0.5 | ● | D |  |  |  |  |  |  | R2 |
| IPH34 | Doing meaningful things |  |  |  |  |  |  |  |  |  |  |  |  | ✓ |
| Personal development |  |  |  |  |  |  |  |  |  |  |  |  |  |  |
| IPH21 | Continue learning |  |  |  |  |  |  |  |  |  |  |  |  | ✓ |
| IPH17 | Wanting to achieve ideals |  |  |  |  |  |  |  |  |  |  |  |  | ✓ |
| IPH13 | Being able to handle changes |  |  |  | FL<0.5 | ● | F |  |  |  |  |  |  | ✓ |
| Cognition |  |  |  |  |  |  |  |  |  |  |  |  |  |  |
| IPH8 | Being able to remember things | 0.7-0.8 | ● | A |  |  |  |  |  |  |  |  |  | R1 |
| IPH9 | Being able to concentrate | 0.7-0.8 | ● | F | FL<0.5 | ● | B |  |  |  |  |  |  | R2 |
| IPH10 | Being able to communicate |  | ● | E |  |  |  |  |  |  |  |  |  | R1 |

IPH; Item number of the original MyPositiveHealth dialogue tool (See Suppl.1)
<sup>1</sup>Results content discussion expressed as A-F <sup>2</sup> to delete or hold an item supported by the measurement properties: ICC; interitem correlations, FL; factor loadings, CA; Cronbach's alphas extracted during exploratory factor analyses.
<sup>2</sup>A to F; A Content is sufficiently reflected in other questions, B Content does not sufficiently match the factor, C Question/Wording not inclusive, D Wording not specific enough, too broad, E Unclear wording, may not be properly understood, F Retained for specific content.

For the factor ‘Cognition’ the content discussion resulted in that only one item was retained. It was accepted by the research team that this factor would not continue to exist as dimension of Positive Health. In total, in **round 1** 12 out of 42 items, originating from each of the six factors, were deleted. For the remaining items (n=30) PCA was applied.

At **round 2** PCA with 30 items resulted in a four-factor structure with explained variance of 60.7% (see Additional file D for factor loadings). Kaiser-Meyer-Olkin (KMO) and Bartlett’s test was statistically significant (0.96; p ≤.001). Factor loadings ranged from 0.369 to 0.780. A new factor with 15 items arose from the former factor ‘Acceptance, meaningfulness and satisfaction with life’ and the factor ‘Social network and societal roles’ of the PH42. Based on the content of these items this new combined factor was renamed by the research team and further called ‘Contentment with life’ (15 items). The other factors were comparable to round 1 (i.e., to the PH42 model), except that the factor Cognition was no longer part of the model. Also, one item; IPH41 about ‘being able to work’, loaded highest, but low (0.495), on the factor ‘Self-management’ instead of the factor ‘Physical health and functioning’. The item about concentration (IPH9), kept from the former factor Cognition, loaded highest, but low (0.369), on the new factor ‘Contentment with life’. Five items had a factor loading (FL) <0.5, and five items loaded also high on another factor (FL>0.32). Of these items, three items were retained based on the content discussion (See Table 1). For example; the items about sleeping pattern (IPH4) and having no pain or complaints (IPH3), both part of the factor ‘Physical health and functioning’, were judged to be specific content that should be held for the measurement tool. For similar reasons item IPH13 (being able to handle changes) was kept. In total, in **round 2** 6 out of 30 items were deleted. In addition, the items selected to delete during round 2 were ranked by the expert team to process the order of item reduction in subsequent PCA. First those items with low factor loadings <0.5 were deleted from the model (in following order; IPH9, IPH35, IPH41). Next those items with also a high factor loading (> 0.32) on another factor were deleted (IPH18, IPH28, IPH24). PCA was executed and checked per deleted item. No changing structures were seen.

In **round 3**, PCA with 24 items resulted in a similar four-factor structure as round 2 with explained variance of 62.4% (see Additional file E for factor loadings). KMO and Bartlett’s test was statistically significant (0.96; p ≤.001). Factor loadings ranged from 0.474 to 0.855. Overall, there was 1 item with a low factor loading (<0.5), and 1 item with factor loadings >0.32 on more than one factor. It concerned the item about sleeping pattern (IPH4), similar to the results of round 2, and the item about asking for help from official institutes (IPH42). Both items were retained because of its specific and relevant content. In this round no items were deleted.

At **round 4** interitem correlations and Cronbach’s alpha (CA) were judged for this four-factor structure with 24 items. For the factor ‘Contentment with life’ 4/11 items were highly correlated (>0.7 but <0.8) and CA was high (0.94). Two additional items were deleted from this factor. There was some doubt about the content of item IPH26 (feeling safe) and its fit among the factor Contentment with life. It was decided to retain this item because it was the only item about this specific subject and considered to be an important aspect of Positive Health. For the factor ‘Physical health and functioning’ two items were highly correlated, but both were kept because of its specific content and good CA of the factor (n=5, CA=0.78). No high interitem correlations nor CA were present among the other factors Self-management (n=5, CA =0.81) and Personal development (n=3, CA=0.74). In total, in **round 4**, 2 out of 24 items were deleted. For the remaining items (n=22) PCA was applied again.

At **round 5,** PCA with 22 items showed similar four-factor structure with explained variance of 62.4% (see Table 2 for factor loadings and Table 3A-D for Interitem correlations). KMO and Bartlett’s test was statistically significant (0.95; p ≤.001). The factor Self-management contained the only item with low FL (0.476). Based on the statistical output and its content no further items were deleted.

**Table 2.** Factor loadings of PH model of 22 items (round 5) (n=1199)

|  | Factor<sup>1</sup> |  |  |  |
| --- | --- | --- | --- | --- |
|  | Contentment with life | Physical fitness | Daily life management | Future perspective |
| IPH32 Having the support of others | **0.800** | -0.139 | 0.042 | 0.057 |
| IPH27 Living conditions | **0.785** | -0.049 | 0.098 | -0.124 |
| IPH29 Social contacts | **0.753** | -0.047 | 0.032 | 0.096 |
| IPH26 Feeling safe | **0.669** | 0.088 | 0.083 | 0.063 |
| IPH16 Being high-spirited | **0.648** | 0.236 | -0.028 | 0.181 |
| IPH19 Accepting life | **0.630** | 0.114 | 0.037 | 0.162 |
| IPH14 Having control | **0.566** | 0.124 | 0.134 | 0.216 |
| IPH12 Accepting yourself | **0.566** | 0.209 | 0.113 | 0.076 |
| IPH34 Doing meaningful things | **0.538** | 0.106 | 0.074 | 0.237 |
| IPH2 Feeling fit | 0.079 | **0.689** | 0.134 | 0.153 |
| IPH6 Physical condition | 0.019 | **0.686** | 0.132 | 0.191 |
| IPH4 Sleeping pattern | 0.385 | **0.667** | -0.101 | -0.193 |
| IPH3 Having physical complaints or pain | -0.145 | **0.560** | 0.052 | 0.153 |
| IPH5 Eating pattern | 0.295 | **0.545** | 0.240 | -0.184 |
| IPH37 Knowing your limitations | 0.033 | 0.026 | **0.855** | -0.062 |
| IPH36 Looking after yourself | -0.087 | 0.123 | **0.793** | 0.021 |
| IPH40 Managing money | 0.101 | -0.036 | **0.781** | -0.081 |
| IPH39 Managing time | -0.018 | 0.067 | **0.768** | 0.054 |
| IPH42 Asking for help | 0.190 | -0.198 | **0.476** | 0.169 |
| IPH21 Continue learning | 0.060 | 0.040 | 0.053 | **0.760** |
| IPH17 Wanting to achieve ideals | 0.153 | 0.147 | -0.061 | **0.705** |
| IPH13 Being able to handle changes | 0.171 | -0.018 | 0.094 | **0.610** |

Extraction Method: Principal Component Analysis. Rotation Method: Oblimin with Kaiser Normalization. Pattern matrix. rotation converged in 8 iterations
<sup>1</sup> During the process of item reduction the names of the factors were Contentment with life, , Physical health and functioning, Self-management and Personal development. Afterwards these were renamed by the research team into Contentment with life, Physical fitness, Daily life management, and Future perspective.

**Table 3A.** Interitem correlation matrix of factor *Contentment with life*^1^ of the 22 item PH model (n=1199)

|  | IPH12 | IPH14 | IPH16 | IPH19 | IPH26 | IPH27 | IPH29 | IPH32 | IPH34 |
| --- | --- | --- | --- | --- | --- | --- | --- | --- | --- |
| IPH12 Accepting yourself | 1.000 | 0.670 | 0.635 | 0.667 | 0.574 | 0.479 | 0.517 | 0.485 | 0.527 |
| IPH14 Having control | 0.670 | 1.000 | 0.669 | 0.664 | 0.639 | 0.545 | 0.591 | 0.516 | 0.583 |
| IPH16 Being high-spirited | 0.635 | 0.669 | 1.000 | 0.670 | 0.597 | 0.558 | 0.590 | 0.548 | 0.628 |
| IPH19 Accepting life | 0.667 | 0.664 | 0.670 | 1.000 | 0.596 | 0.492 | 0.512 | 0.506 | 0.524 |
| IPH26 Feeling safe | 0.574 | 0.639 | 0.597 | 0.596 | 1.000 | 0.605 | 0.523 | 0.552 | 0.521 |
| IPH27 Living conditions | 0.479 | 0.545 | 0.558 | 0.492 | 0.605 | 1.000 | 0.509 | 0.582 | 0.485 |
| IPH29 Social contacts | 0.517 | 0.591 | 0.590 | 0.512 | 0.523 | 0.509 | 1.000 | 0.695 | 0.607 |
| IPH32 Having support of others | 0.485 | 0.516 | 0.548 | 0.506 | 0.552 | 0.582 | 0.695 | 1.000 | 0.557 |
| IPH34 Doing meaningful things | 0.527 | 0.583 | 0.628 | 0.524 | 0.521 | 0.485 | 0.607 | 0.557 | 1.000 |

**Table 3B.**
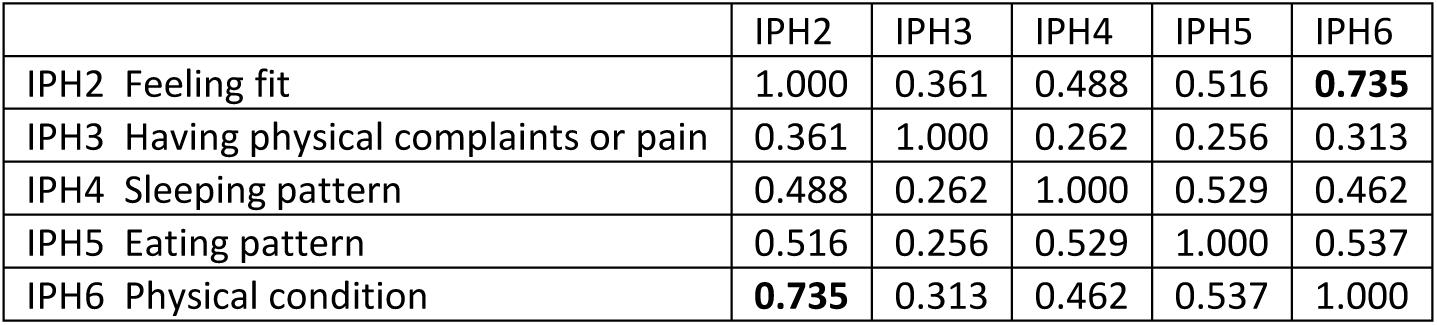
Interitem correlation matrix of factor *Physical fitness*^1^ of the 22 item PH model (n=1199)

**Table 3C.**
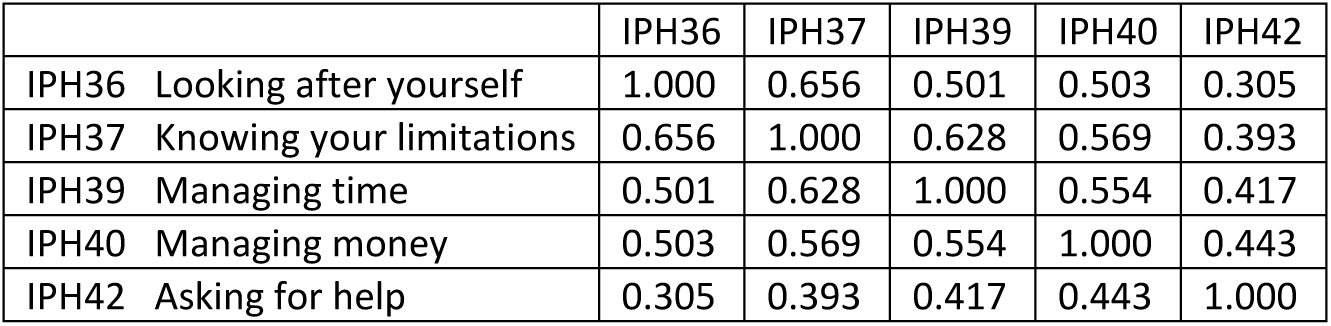
Interitem correlation matrix of factor *Daily life management*^1^ of the 22 item PH model (n=1199)

|  | IPH36 | IPH37 | IPH39 | IPH40 | IPH42 |
| --- | --- | --- | --- | --- | --- |
| IPH36 Looking after yourself | 1.000 | 0.656 | 0.501 | 0.503 | 0.305 |
| IPH37 Knowing your limitations | 0.656 | 1.000 | 0.628 | 0.569 | 0.393 |
| IPH39 Managing time | 0.501 | 0.628 | 1.000 | 0.554 | 0.417 |
| IPH40 Managing money | 0.503 | 0.569 | 0.554 | 1.000 | 0.443 |
| IPH42 Asking for help | 0.305 | 0.393 | 0.417 | 0.443 | 1.000 |

**Table 3D.** Interitem correlation matrix of factor *Future perspective*^1^ of the 22 item PH model (n=1199)

|  | IPH13 | IPH17 | IPH21 |
| --- | --- | --- | --- |
| IPH13 Being able to handle changes | 1.000 | 0.449 | 0.483 |
| IPH17 Wanting to achieve ideals | 0.449 | 1.000 | 0.534 |
| IPH21 Continue learning | 0.483 | 0.534 | 1.000 |

<sup>1</sup> During the process of item reduction the names of the factors were Contentment with life, , Physical health and functioning, Self-management and Personal development. Afterwards these were renamed by the research team into Contentment with life, Physical fitness, Daily life management, and Future perspective.

In summary, through the 5 rounds of item reduction evaluation and discussions, 20 out of 42 items were deleted resulting in a short self-reported questionnaire to measure Positive Health consisting of four dimensions and 22 items, hereafter called the PH22. The dimensions were renamed by the research team into 1) Physical fitness, 2) Contentment with life, 3) Daily life management, and 4) Future perspective (see Table 4).

**Table 4.** The 22 item self-reported Positive Health questionnaire (PH22)

| Item number of the dialogue tool | Item |
| --- | --- |
| Physical fitness |  |
| IPH2 | I feel fit |
| IPH3 | I have no physical complaints or pain |
| IPH4 | I sleep well |
| IPH5 | I eat well |
| IPH6 | I recover quickly after exercise, such as sports |
| Contentment with life |  |
| IPH12 | I accept myself for who I am |
| IPH14 | I feel in control of my life |
| IPH16 | In the mornings. I look forward to the day ahead |
| IPH19 | I accept life as it comes |
| IPH26 | I feel safe |
| IPH27 | I am content with where and with whom I live |
| IPH29 | I am in good contact with other people |
| IPH32 | I have people who support me when I need it |
| IPH34 | I consider my job or other activities to be meaningful |
| Daily life management |  |
| IPH36 | I am well capable of looking after myself. for example with regard to personal hygiene. getting dressed. shopping. cooking |
| IPH37 | I know my limitations |
| IPH39 | I am well capable of planning my day |
| IPH40 | I am well capable of managing the money that I have each month |
| IPH42 | I know how to apply for benefits or getting assistance from official agencies when necessary |
| Future perspective |  |
| IPH13 | I look for solutions to change difficult situations |
| IPH17 | I have ideals that I would like to achieve |
| IPH21 | I want to continue learning throughout my life |

It was accepted for the PH22 in favour of keeping specific content that; 1) the factor ‘Contentment with life’ had high CA (0.92), 2) the factor ‘Physical fitness’ contained two highly correlated items but with an adequate CA of 0.78, and 3) the factor ‘Daily life management’ contained an item with low FL (also an adequate CA of 0.81).

### Cross-validation

The four-factor structure of the PH22 had an acceptable fit in first and second order CFA; 1) significant X^2^ (p≤0.001), CFI of 0.902, RMSEA of 0.079 with a 90% confidence interval of 0.076 to 0.082, and SMSR of 0.047, and 2) significant X^2^ (p≤0.001), CFI of 0.901, RMSEA of 0.079 with a 90% confidence interval of 0.075 to 0.0782, and SMSR of 0.047, respectively.

### Scores of the developed short Positive Health questionnaire

The scores of the PH22 were interpreted normally distributed but with slightly more outliers for the lower scores and higher frequency of scores around the mean, which was especially seen for the scores of the factor ‘Daily life management’. No floor or ceiling effects were present (see Table 5).

**Table 5.**
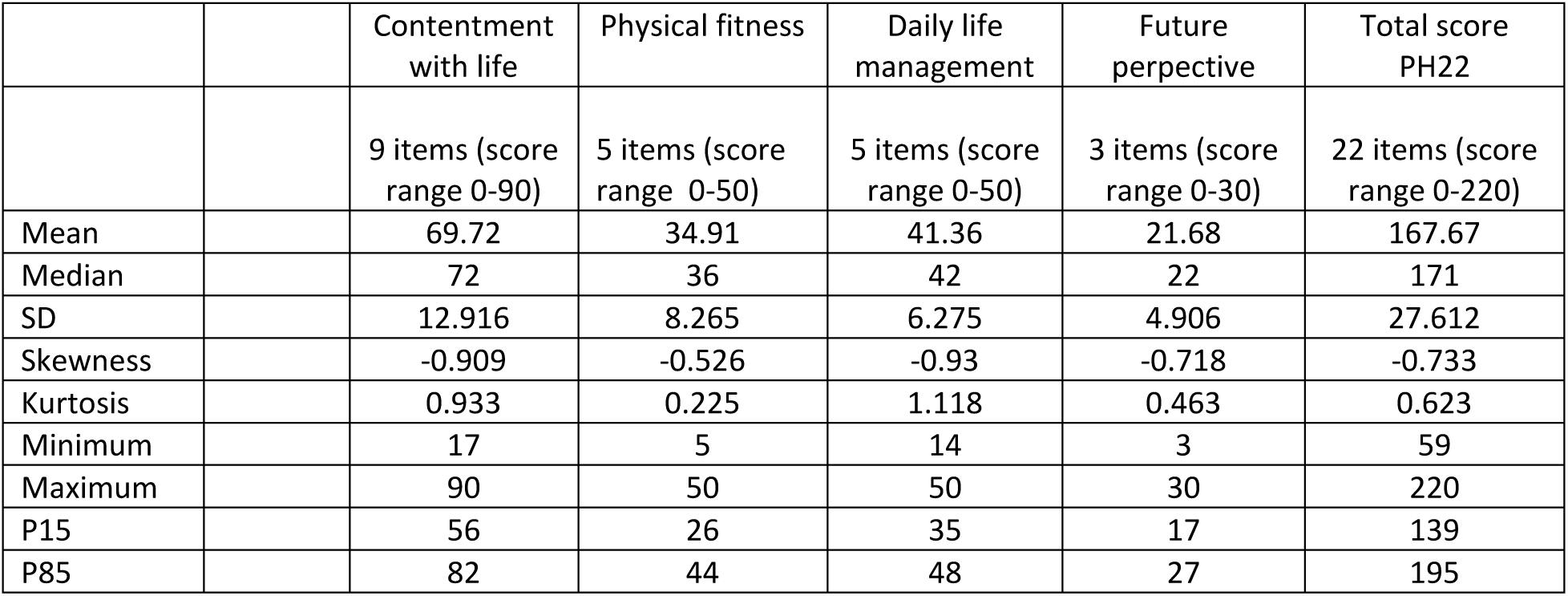
Descriptive statistics of the PH22 scores (n=1258)

|  |  | Contentment with life | Physical fitness | Daily life management | Future perspective | Total score PH22 |
| --- | --- | --- | --- | --- | --- | --- |
|  |  | 9 items (score range 0-90) | 5 items (score range 0-50) | 5 items (score range 0-50) | 3 items (score range 0-30) | 22 items (score range 0-220) |
| Mean |  | 69.72 | 34.91 | 41.36 | 21.68 | 167.67 |
| Median |  | 72 | 36 | 42 | 22 | 171 |
| SD |  | 12.916 | 8.265 | 6.275 | 4.906 | 27.612 |
| Skewness |  | -0.909 | -0.526 | -0.93 | -0.718 | -0.733 |
| Kurtosis |  | 0.933 | 0.225 | 1.118 | 0.463 | 0.623 |
| Minimum |  | 17 | 5 | 14 | 3 | 59 |
| Maximum |  | 90 | 50 | 50 | 30 | 220 |
| P15 |  | 56 | 26 | 35 | 17 | 139 |
| P85 |  | 82 | 44 | 48 | 27 | 195 |

## DISCUSSION

In this study a relatively short questionnaire to measure self-reported Positive Health was composed and cross-validated among a general (Dutch) population. The questionnaire contains 22 items stemming from the original My Positive Health (MPH) dialogue tool with 42 items. Structural validity and internal consistency were satisfactory, supporting the use of this questionnaire for evaluative purposes in scientific or policy research. This questionnaire is called the PH22.

The different methodological approaches of item reduction for the PH17(9) and PH22 resulted in a different set of items and measurement properties. Contrary to the development of the PH17, during the development of the PH22, the approach by De Vet et al.(12) was used for item reduction, which includes content discussion and judgement of internal consistency next to highest factor loadings. First, these steps are considered essential to the item reduction process to avoid withdrawing relevant items. Second, retaining items with the highest factor loadings per factor without the other steps can lead to overlap, i.e. the answer to one question predicts the answer to the second, thus providing information as if it were merely one item. Overall, the approach by De Vet et al.(12) most likely improves a questionnaire’s discriminative ability, which means that a tool is better able to generate different scores for populations with different levels of Positive Health. This is considered an essential condition for a measurement instrument, particularly for instruments aiming to evaluate interventions or follow cohorts. The too high internal consistency found for at least parts of the PH17 dimensions might be a consequence of this. Looking at the PH17, internal inconsistency was high for almost all dimensions, especially related to the low number of items per factor (2-3 items; Cronbach’s alpha (CA); 0.90, 0.89, 0.77, 0.93, 0.89, 0.84). More items result in higher CA by definition. For the PH22, the dimension ‘Contentment with life’ also had too high internal consistency (CA=0.92), but the factor also consisted of nine items, what might (partly) explain the high CA. The other dimensions of the PH22 showed good internal consistency, with CA ranging from 0.74 to 0.81. Finally, both PH17(9) and PH22 development started with the 42 items of the MPH dialogue, but the different methodological approaches resulted in other sets of items; only eight items corresponded. When comparing the PH22 to the PH42(11), its internal consistency and user-friendliness improved because of fewer items, at the expense of only a bit less explained variance (62% and 68%, respectively).

We presumed the 42 items of the MPH to be a content-valid basis to compose a measurement instrument, reflecting ‘health from the perspective of patients and citizens’ as assessed by Huber et al(5). The items of the MPH tool were formulated based on health indicators emerged from a large concept elicitation interview study among various stakeholders including patients and citizens(5), generating a solid basis for its content. In the meantime, studies showed that scores from the PH17 and PH42 correlated with constructs like quality of life, resilience and recovery (10,11,24) and with level of education and healthcare use (24).

Moreover, the MPH was shown by various users as a relevant and comprehensible dialogue tool(3). We followed an inductive approach towards the development of the PH22. Thereby, four dimensions emerged which we named; ‘Physical fitness’, ‘Contentment with life’, ‘Daily life management’ and ‘Future perspective’ aligning with the core elements of the dynamic concept of (positive) health by Huber et al.(2,5).

During the development of the dynamic concept by Huber et al.(2) and during its elaboration into Positive Health(5), a deliberate choice was made to strive for an open concept instead of a more demarcated definition. Nevertheless, when creating a measurement instrument, it is important to establish a clear construct(25). It should be noted that no widely agreed construct for Positive Health exists so far (25,26). As described above, in this study we chose the construct for the measurement tool to reflect the original concept of health by Huber et al (2) ‘Health as the ability to adapt and to self-manage, in the face of social, physical and emotional challenges’. This concept closely fits a recently proposed description of positive health: ‘reserve in capacities’(26). Recently, another Dutch research group published the 32-item Context-sensitive Positive Health Questionnaire (CPHQ)(27). This measurement tool aligns the concept of Positive Health with the ‘Capability Approach’ (28). Accordingly, they formulated the following construct definition for their measurement tool: “The extent to which one is capable to adapt and to thrive given one’s physical, mental, social and contextual opportunities”. As a result, the CPHQ included more context-related items than the PH22, such as items about feeling disadvantaged because of sexuality or cultural background or feeling represented by politics. Nevertheless, the PH22 and CPHQ also overlap, both including capabilities and functionings (beings and doings). For the methodological process of item reduction towards the 32 item CPHQ, similar as were for the PH17, the three items with highest factor loadings (>0.4 without cross-loadings) were leading, possibly hampering its discriminant validity. Last, contrary to the CPHQ, the PH22 consists only of original items from the MPH to keep recognizability with the Positive Health approach in practice. As ‘Positive Health’ is a novice approach, the discussion as to which construct or theoretical framework approximates best should continue. Moreover, Van Druten et al.(15) pointed out that conceptualization of health is person– and context-dependent, which necessitates the existence of various constructs. Therefore, different definitions and theoretical frameworks, such as Positive Health, Reserve Capacity Model(29) or Capability Approach(28), should exist side by side. At the moment the CPHQ is being further developed and assessed(30). One part of the research consists of comprehensive focus groups with various stakeholders discussing and prioritizing items anew with the aim to shorten the questionnaire and resulting in a broad supported instrument to assess the broad concept of health. It is of interest to explore how these instruments can supplement each other, or in other words, which instrument serves which aim and context best. Future choices of which tool to use should not only depend on the measurement properties and usability of each tool but also on which construct definition is preferred as the outcome to measure (8,15).

The PH22 scores can add to evaluate positive health or patient centered interventions. Prior to the actual use of the PH22 as a measuring tool in evaluative research, it is essential to explore its test-retest reliability and responsiveness for change. Further research has to explore this so that differences in scores can be correctly interpreted. Last, it should be emphasized that the PH22 is not meant for dialogue purposes.

Specifically, for that aim the MPH dialogue tool was developed; to guide the conversation about someone’s Positive Health and reflect on someone’s personalized (positive) health-related goals over time in his or her specific context.

## Data Availability

The original dataset(s) supporting the conclusions of this article is(are) available from https://www.lissdata.nl/access-data upon request for researchers and policymakers.

## CONCLUSIONS

In this study a comprehensive methodological approach was applied using both content discussion and statistical output aiming to develop a content valid measurement tool for evaluative purposes in scientific or policy research at positive health or patient centered interventions assessing self-reported Positive Health. A relatively short questionnaire containing 22 items distributed over four dimensions, the PH22, was developed and cross-validated among a general (Dutch) population. This study supports its structural validity. To apply this questionnaire in evaluative research its test-retest reliability should be explored first, followed by responsiveness for change. Future research has to assess this.

### Additional material

Additional file (.pdf); A. Items of the My Positive Health dialogue tool (MPH), B. Factor loadings of model with 42 MPH items; PH42, C1-6. Interitem Correlations of factors PH42, D-E. Factor loadings with 30-item and 24 PH model (round 2 and 3).

## DECLARATIONS

### Ethical approval and consent to participate

The study was conducted in accordance with current public regulations, laws, and the principles of the Declaration of Helsinki. Informed consent was given by each participant to be included as a LISS-panel member. For more information see: https://www.lisspanel.nl/ethics. The Medical Ethics Committee of Brabant (Tilburg, the Netherlands) reviewed this study and declared that the Medical Research Involving Human Subjects Act (WMO) did not apply to this study (study number NW2024-15).

### Consent for publication

Not applicable

### Author contributions

LNvV wrote the protocol and manuscript, and conducted the statistical analyses. VvD contributed to the first part of the statistical analysis. BvdZ supervised the research, statistical analyses and the writing process. LNvV, BvdZ, MM and MvV participated at the research meetings concerning the item reduction process; content discussion and interpretation of the statistical output. MM and MvV contributed equally. All author’s reviewed and approved the final manuscript.

## Acknowledgements

We thank Miriam de Kleijn from the Institute for Positive Health for her participation at our discussion on forehand about the aim of the measurement tool, and later on, on (re)naming the factors of the PH22. We also thank all panel members for their contribution.

## List of abbreviations

CA: Cronbach’s alpha
CFA: Confirmatory factor analysis
CFI: Comparative fit index
COSMIN: Reporting Guideline
CPHQ: Context-sensitive Positive Health Questionnaire
PCA: Principal component analysis
FL: Factor loading
IIC: Inter-item correlation
IPH: Item number from the MPH dialogue tool
KMO: Kaiser-Meyer-Olkin
LISS panel: Longitudinal Internet studies for the Social Sciences – panel
METC: Medical ethical review board (Medisch ethische toetsingscommissie)
ML: Maximum likelihood
MPH: My Positive Health dialogue tool
PH17: Positive Health measurement scale with 17 items
PH22: Positive Health measurement scale with 22 items
PH42: Positive Health measurement scale with 42 items
PROM: Patient-reported outcome measures
RMSEA: Root mean square error of approximation
STMR: Standardized root mean square residual

