## Additional file A-E for "Development and Cross-Validation of a Short Questionnaire to Evaluate Self-Reported Positive Health; A Cross Sectional Panel Study of Structural Validity Among a General Dutch Population"

**A.** Items of the My Positive Health dialogue tool (MPH)

| Item number of the dialogue tool | Item |
|---|---|
| BF1 | I feel healthy |
| BF2 | I feel fit |
| BF3 | I have no physical complaints or pain |
| BF4 | I sleep well |
| BF5 | I eat well |
| BF6 | I recover quickly after exercise, such as sports. |
| BF7 | I find it easy to move, such as going up and down stairs, walking or cycling |
| MW8 | I am good at remembering things |
| MW9 | I am able to concentrate |
| MW10 | I am able to see, hear, talk and read |
| MW11 | I feel cheerful |
| MW12 | I accept myself for who I am |
| MW13 | I look for solutions to change difficult situations |
| MW14 | I feel in control of my life |
| MF15 | I have a meaningful life |
| MF16 | In the mornings, I look forward to the day ahead |
| MF17 | I have ideals that I would like to achieve |
| MF18 | I feel confident about my own future |
| MF19 | I accept life as it comes |
| MF20 | I am grateful for what life offers me |
| MF21 | I want to continue learning throughout my life |
| QL22 | I enjoy my life |
| QL23 | I am happy |
| QL24 | I feel good |
| QL25 | I feel my life is well-balanced |
| QL26 | I feel safe |
| QL27 | I am content with where and with whom I live |
| QL28 | I have enough money to pay my bills |
| SP29 | I am in good contact with other people |
| SP30 | Other people take me seriously |
| SP31 | I have people with whom I can do fun things with |
| SP32 | I have people who support me when I need it |
| SP33 | I feel that I 'belong' in my environment |
| SP34 | I consider my job or other activities to be meaningful |
| SP35 | I am interested in what happens in society |
| DF36 | I am well capable of looking after myself, for example with regard to personal hygiene, getting dressed, shopping, cooking |
| DF37 | I know my limitations |
| DF38 | I know how I can look after my own health |
| DF39 | I am well capable of planning my day |
| DF40 | I am well capable of managing the money that I have each month |
| DF41 | I am able to work in a job or do voluntary work |
| DF42 | I know how to apply for benefits or getting assistance from official agencies when necessary |

BF: bodily functions, MW: mental wellbeing, MF: meaningfulness, QL: quality of life, SP: social and societal participation, DF: daily functioning (https://vragenlijsten.mijnpositievegezondheid.nl/adults-en)

**B.** Factor loadings of model with 42 MPH items; PH42 (n=1199)[1]

| Item number of the My Positive Health dialogue tool (MPH) (Expressed as IPH) | Acceptance. meaningfulness and satisfaction with life | Physical health and functioning | Self-management | Social network and societal roles | Personal development | Cognition |
|---|---|---|---|---|---|---|
| IPH25 Feeling well-balanced | **0.751** | 0.064 | 0.129 | 0.053 | 0.047 | 0.036 |
| IPH24 Feeling good | **0.708** | 0.235 | -0.009 | 0.097 | 0.051 | 0.005 |
| IPH23 Being happy | **0.699** | 0.082 | -0.004 | 0.260 | 0.034 | -0.023 |
| IPH22 Enjoyment | **0.676** | 0.086 | 0.021 | 0.253 | 0.060 | -0.013 |
| IPH16 Being high-spirited | **0.673** | 0.105 | 0.046 | 0.129 | 0.102 | 0.033 |
| IPH11 Being cheerful | **0.653** | 0.159 | -0.040 | 0.154 | 0.089 | 0.102 |
| IPH19 Accepting life | **0.645** | -0.039 | 0.165 | 0.048 | 0.135 | 0.087 |
| IPH20 Being grateful | **0.624** | 0.008 | 0.114 | 0.135 | 0.160 | 0.002 |
| IPH15 Having a meaningful life | **0.573** | 0.049 | 0.045 | 0.249 | 0.183 | 0.005 |
| IPH12 Accepting yourself | **0.572** | 0.037 | 0.206 | 0.054 | 0.043 | 0.145 |
| IPH18 Feeling confident about own future | **0.558** | 0.161 | 0.033 | 0.093 | 0.344 | -0.019 |
| IPH14 Having control | **0.534** | 0.026 | 0.201 | 0.105 | 0.163 | 0.123 |
| IPH26 Feeling safe | **0.393** | 0.074 | 0.185 | 0.256 | 0.028 | 0.078 |
| IPH7 Exercise | -0.107 | **0.877** | 0.061 | 0.049 | 0.109 | -0.118 |
| IPH6 Physical condition | 0.069 | **0.783** | 0.046 | 0.021 | 0.048 | -0.013 |
| IPH2 Feeling fit | 0.206 | **0.781** | 0.052 | -0.039 | -0.002 | 0.003 |
| IPH1 Feeling healthy | 0.195 | **0.769** | 0.027 | -0.032 | 0.016 | 0.028 |
| IPH41 Being able to work | -0.089 | **0.526** | 0.291 | 0.058 | 0.266 | -0.223 |
| IPH3 Having physical complaints or pain | -0.031 | **0.450** | -0.097 | 0.062 | -0.018 | 0.193 |
| IPH4 Sleeping pattern | 0.395 | **0.422** | -0.061 | 0.022 | -0.276 | 0.254 |
| IPH5 Eating pattern | 0.241 | **0.364** | 0.248 | 0.087 | -0.250 | 0.197 |
| IPH40 Managing money | 0.147 | -0.036 | **0.828** | 0.001 | -0.069 | -0.070 |
| IPH37 Knowing your limitations | 0.001 | 0.014 | **0.754** | 0.017 | 0.011 | 0.233 |
| IPH38 Knowledge of health | -0.022 | 0.157 | **0.651** | 0.088 | 0.019 | 0.201 |
| IPH39 Managing time | 0.132 | 0.001 | **0.634** | -0.062 | 0.073 | 0.238 |
| IPH36 Looking after yourself | -0.171 | 0.290 | **0.633** | 0.052 | 0.092 | 0.065 |
| IPH28 Having enough money | 0.219 | 0.035 | **0.598** | 0.147 | -0.121 | -0.231 |
| IPH42 Asking for help | 0.065 | -0.044 | **0.452** | 0.178 | 0.175 | -0.154 |
| IPH32 Having the support of others | -0.014 | -0.013 | -0.036 | **0.939** | -0.038 | 0.015 |
| IPH31 Doing fun things together | 0.043 | 0.070 | -0.070 | **0.899** | -0.014 | -0.043 |
| IPH33 Belonging | 0.099 | -0.015 | 0.018 | **0.864** | -0.070 | -0.028 |
| IPH30 Being taken seriously | -0.023 | -0.021 | 0.047 | **0.792** | 0.081 | 0.092 |
| IPH29 Social contacts | 0.111 | 0.013 | -0.038 | **0.786** | 0.013 | 0.051 |
| IPH27 Living conditions | 0.335 | -0.044 | 0.224 | **0.423** | -0.140 | -0.017 |
| IPH35 Being interested in society | 0.011 | -0.002 | 0.181 | **0.418** | 0.263 | -0.011 |
| IPH34 Doing meaningful things | 0.228 | 0.212 | 0.103 | **0.401** | 0.181 | -0.169 |
| IPH21 Continue learning | 0.182 | 0.118 | 0.020 | 0.002 | **0.660** | 0.017 |
| IPH17 Wanting to achieve ideals | 0.310 | 0.161 | -0.115 | -0.008 | **0.643** | 0.051 |

Extraction Method: Principal Component Analysis. Rotation Method: Oblimin with Kaiser Normalization. Pattern matrix, rotation converged in 14 iterations.

1. van Druten VP, Metz MJ, Mathijssen JJP, van Vliet M, Rudd B, de Vries E, Nahar -van Venrooij L.M.W. Measuring positive health using the My Positive Health (MPH) and Individual Recovery Outcomes Counter (I.ROC) dialogue tools: a panel study on measurement properties in a representative general Dutch population. Applied Research in Quality of Life. Vol.21, 2024. https://doi.org/10.1007/s11482-024-10356-3

**C1.** Interitem Correlations of factor *Acceptation, meaningfulness and satisfaction with life* of 42-item model; PH42[1]

| | IPH23 Being happy | IPH22 Enjoyment | IPH25 Feeling well-balanced | IPH24 Feeling good | IPH16 Being high-spirited | IPH11 Being cheerful | IPH15 Having a meaningful life | IPH20 Being grateful | IPH18 Feeling confident about own future | IPH19 Accepting life | IPH14 Having control | IPH12 Accepting yourself | IPH26 Feeling safe |
|---|---|---|---|---|---|---|---|---|---|---|---|---|---|
| IPH23 Being happy | 1.000 | 0.893 | 0.811 | 0.844 | 0.784 | 0.815 | 0.769 | 0.756 | 0.746 | 0.664 | 0.699 | 0.652 | 0.646 |
| IPH22 Enjoyment | 0.893 | 1.000 | 0.804 | 0.842 | 0.793 | 0.822 | 0.785 | 0.769 | 0.757 | 0.678 | 0.705 | 0.669 | 0.658 |
| IPH25 Feeling well-balanced | 0.811 | 0.804 | 1.000 | 0.831 | 0.764 | 0.755 | 0.733 | 0.702 | 0.723 | 0.689 | 0.709 | 0.670 | 0.640 |
| IPH24 Feeling good | 0.844 | 0.842 | 0.831 | 1.000 | 0.784 | 0.827 | 0.726 | 0.682 | 0.724 | 0.657 | 0.680 | 0.687 | 0.648 |
| IPH16 Being high-spirited | 0.784 | 0.793 | 0.764 | 0.784 | 1.000 | 0.773 | 0.770 | 0.697 | 0.738 | 0.670 | 0.669 | 0.635 | 0.597 |
| IPH11 Being cheerful | 0.815 | 0.822 | 0.755 | 0.827 | 0.773 | 1.000 | 0.721 | 0.691 | 0.723 | 0.654 | 0.676 | 0.653 | 0.634 |
| IPH15 Having a meaningful life | 0.769 | 0.785 | 0.733 | 0.726 | 0.770 | 0.721 | 1.000 | 0.716 | 0.750 | 0.663 | 0.710 | 0.631 | 0.614 |
| IPH20 Being grateful | 0.756 | 0.769 | 0.702 | 0.682 | 0.697 | 0.691 | 0.716 | 1.000 | 0.712 | 0.709 | 0.653 | 0.647 | 0.613 |
| IPH18 Feeling confident about own future | 0.746 | 0.757 | 0.723 | 0.724 | 0.738 | 0.723 | 0.750 | 0.712 | 1.000 | 0.697 | 0.715 | 0.640 | 0.623 |
| IPH19 Accepting life | 0.664 | 0.678 | 0.689 | 0.657 | 0.670 | 0.654 | 0.663 | 0.709 | 0.697 | 1.000 | 0.664 | 0.667 | 0.596 |
| IPH14 Having control | 0.699 | 0.705 | 0.709 | 0.680 | 0.669 | 0.676 | 0.710 | 0.653 | 0.715 | 0.664 | 1.000 | 0.670 | 0.639 |
| IPH12 Accepting yourself | 0.652 | 0.669 | 0.670 | 0.687 | 0.635 | 0.653 | 0.631 | 0.647 | 0.640 | 0.667 | 0.670 | 1.000 | 0.574 |
| IPH26 Feeling safe | 0.646 | 0.658 | 0.640 | 0.648 | 0.597 | 0.634 | 0.614 | 0.613 | 0.623 | 0.596 | 0.639 | 0.574 | 1.000 |

**C2.** Interitem Correlations between items of factor *Physical health and functioning* of 42-item model; PH42[1]

| | IPH2 Feeling fit | IPH7 Exercise | IPH1 Feeling healthy | IPH6 Physical condition | IPH41 Being able to work | IPH4 Sleeping pattern | IPH5 Eating pattern | IPH3 Having complaints or pain |
|---|---|---|---|---|---|---|---|---|
| IPH2 Feeling fit | 1.000 | 0.704 | 0.845 | 0.735 | 0.490 | 0.488 | 0.516 | 0.361 |
| IPH7 Exercise | 0.704 | 1.000 | 0.682 | 0.735 | 0.548 | 0.395 | 0.435 | 0.312 |
| IPH1 Feeling healthy | 0.845 | 0.682 | 1.000 | 0.674 | 0.518 | 0.488 | 0.476 | 0.348 |
| IPH6 Physical condition | 0.735 | 0.735 | 0.674 | 1.000 | 0.477 | 0.462 | 0.537 | 0.313 |
| IPH41 Being able to work | 0.490 | 0.548 | 0.518 | 0.477 | 1.000 | 0.283 | 0.294 | 0.184 |
| IPH4 Sleeping pattern | 0.488 | 0.395 | 0.488 | 0.462 | 0.283 | 1.000 | 0.529 | 0.262 |
| IPH5 Eating pattern | 0.516 | 0.435 | 0.476 | 0.537 | 0.294 | 0.529 | 1.000 | 0.256 |
| IPH3 Having complaints or pain | 0.361 | 0.312 | 0.348 | 0.313 | 0.184 | 0.262 | 0.256 | 1.000 |

**C3.** Interitem Correlations between items of factor *Self-management* of 42-item model; PH42[1]

| | IPH40 Managing money | IPH37 Knowing your limitations | IPH38 Knowledge of health | IPH36 Looking after yourself | IPH39 Managing time | IPH28 Having enough money | IPH42 Asking for help |
|---|---|---|---|---|---|---|---|
| IPH40 Managing money | 1.000 | 0.569 | 0.570 | 0.503 | 0.554 | 0.690 | 0.443 |
| IPH37 Knowing your limitations | 0.569 | 1.000 | 0.779 | 0.656 | 0.628 | 0.403 | 0.393 |
| IPH38 Knowledge of health | 0.570 | 0.779 | 1.000 | 0.666 | 0.602 | 0.413 | 0.404 |
| IPH36 Looking after yourself | 0.503 | 0.656 | 0.666 | 1.000 | 0.501 | 0.359 | 0.305 |
| IPH39 Managing time | 0.554 | 0.628 | 0.602 | 0.501 | 1.000 | 0.388 | 0.417 |
| IPH28 Having enough money | 0.690 | 0.403 | 0.413 | 0.359 | 0.388 | 1.000 | 0.379 |
| IPH42 Asking for help | 0.443 | 0.393 | 0.404 | 0.305 | 0.417 | 0.379 | 1.000 |

**C4.** Interitem Correlations between items of factor *Social network and societal roles* of 42-item model; PH42[1]

| | IPH31 Doing fun things together | IPH32 Having the support of others | IPH33 Belonging | IPH29 Social contacts | IPH30 Being taken seriously | IPH34 Doing meaningful things | IPH27 Living conditions | IPH35 Being interested in society |
|---|---|---|---|---|---|---|---|---|
| IPH31 Doing fun things together | 1.000 | 0.822 | 0.774 | 0.734 | 0.694 | 0.598 | 0.538 | 0.492 |
| IPH32 Having the support of others | 0.822 | 1.000 | 0.779 | 0.695 | 0.678 | 0.557 | 0.582 | 0.467 |
| IPH33 Belonging | 0.774 | 0.779 | 1.000 | 0.743 | 0.711 | 0.621 | 0.569 | 0.523 |
| IPH29 Social contacts | 0.734 | 0.695 | 0.743 | 1.000 | 0.724 | 0.607 | 0.509 | 0.481 |
| IPH30 Being taken seriously | 0.694 | 0.678 | 0.711 | 0.724 | 1.000 | 0.561 | 0.533 | 0.540 |
| IPH34 Doing meaningful things | 0.598 | 0.557 | 0.621 | 0.607 | 0.561 | 1.000 | 0.485 | 0.521 |
| IPH27 Living conditions | 0.538 | 0.582 | 0.569 | 0.509 | 0.533 | 0.485 | 1.000 | 0.391 |
| IPH35 Being interested in society | 0.492 | 0.467 | 0.523 | 0.481 | 0.540 | 0.521 | 0.391 | 1.000 |

**C5.** Interitem Correlations between items of factor *Personal development* of 42-item model; PH42[1]

| | IPH21 Continue learning | IPH17 Wanting to achieve ideals | IPH13 Being able to handle changes |
|---|---|---|---|
| IPH21 Continue learning | 1.000 | 0.534 | 0.483 |
| IPH17 Wanting to achieve ideals | 0.534 | 1.000 | 0.449 |
| IPH13 Being able to handle changes | 0.483 | 0.449 | 1.000 |

**C6.** Interitem Correlations between items of factor *Cognition* of 42-item model; PH42[1]

| | IPH8 Being able to remember things | IPH9 Being able to concentrate | IPH10 Being able to communicate |
|---|---|---|---|
| IPH8 Being able to remember things | 1.000 | 0.768 | 0.477 |
| IPH9 Being able to concentrate | 0.768 | 1.000 | 0.452 |
| IPH10 Being able to communicate | 0.477 | 0.452 | 1.000 |

1. van Druten VP, Metz MJ, Mathijssen JJP, van Vliet M, Rudd B, de Vries E, Nahar-van Venrooij L.M.W.. Measuring positive health using the My Positive Health (MPH) and Individual Recovery Outcomes Counter (I.ROC) dialogue tools: a panel study on measurement properties in a representative general Dutch population. Applied Research in Quality of Life. Vol.21, 2024. https://doi.org/10.1007/s11482-024-10356-3

**D.** Factor loadings of 30-item PH model (round 2) (n=1199)

| | Factor[1] | | | |
|---|---|---|---|---|
| | Contentment with life | Daily life management | Physical fitness | Future perspective |
| IPH32 Having the support of others | **0.757** | 0.116 | -0.196 | 0.045 |
| IPH20 Being grateful | **0.731** | -0.027 | 0.114 | 0.129 |
| IPH29 Social contacts | **0.730** | 0.097 | -0.107 | 0.097 |
| IPH19 Accepting life | **0.729** | -0.022 | 0.136 | 0.064 |
| IPH16 Being high-spirited | **0.712** | -0.049 | 0.246 | 0.116 |
| IPH27 Living conditions | **0.710** | 0.168 | -0.046 | -0.157 |
| IPH24 Feeling good | **0.692** | -0.087 | 0.344 | 0.106 |
| IPH30 Being taken seriously | **0.674** | 0.194 | -0.183 | 0.122 |
| IPH26 Feeling safe | **0.673** | 0.100 | 0.103 | 0.002 |
| IPH12 Accepting yourself | **0.666** | 0.045 | 0.217 | -0.011 |
| IPH14 Having control | **0.629** | 0.111 | 0.156 | 0.121 |
| IPH18 Feeling confident about own future | **0.610** | -0.038 | 0.197 | 0.350 |
| IPH34 Doing meaningful things | **0.472** | 0.187 | 0.079 | 0.260 |
| IPH35 Being interested in society | **0.399** | 0.263 | -0.139 | 0.256 |
| IPH9 Being able to concentrate | **0.369** | 0.218 | 0.268 | 0.041 |
| IPH40 Managing money | 0.154 | **0.780** | -0.031 | -0.158 |
| IPH37 Knowing your limitations | 0.067 | **0.762** | 0.073 | -0.049 |
| IPH36 Looking after yourself | -0.107 | **0.750** | 0.161 | 0.096 |
| IPH39 Managing time | 0.041 | **0.693** | 0.118 | 0.033 |
| IPH28 Having enough money | 0.345 | **0.535** | -0.043 | -0.178 |
| IPH42 Asking for help | 0.166 | **0.502** | -0.174 | 0.174 |
| IPH41 Being able to work | -0.161 | **0.495** | 0.257 | 0.388 |
| IPH2 Feeling fit | 0.106 | 0.161 | **0.674** | 0.155 |
| IPH6 Physical condition | 0.031 | 0.193 | **0.648** | 0.197 |
| IPH4 Sleeping pattern | 0.364 | -0.071 | **0.644** | -0.204 |
| IPH5 Eating pattern | 0.316 | 0.238 | **0.511** | -0.222 |
| IPH3 Having physical complaints or pain | -0.052 | 0.064 | **0.485** | 0.107 |
| IPH17 Wanting to achieve ideals | 0.269 | -0.083 | 0.152 | **0.663** |
| IPH21 Continue learning | 0.230 | 0.032 | 0.032 | **0.640** |
| IPH13 Being able to handle changes | 0.289 | 0.101 | -0.016 | **0.465** |

Extraction Method: Principal Component Analysis; Rotation Method: Oblimin with Kaiser Normalization; Pattern matrix, rotation converged in 16 iterations

[1] During the process of item reduction the names of the factors were Contentment with life, Self-management, Physical health and functioning, and Personal development. Afterwards these were renamed by the research team into Contentment with life, Daily life management, Physical fitness, and Future perspective.

**E.** Factor loadings of 24-item PH model (round 3) (n=1199)

| | Factor[1] | | | |
|---|---|---|---|---|
| | Contentment with life | Physical fitness | Daily life management | Future perspective |
| IPH32 Having the support of others | **0.846** | -0.136 | 0.023 | 0.009 |
| IPH29 Social contacts | **0.806** | -0.055 | 0.014 | 0.051 |
| IPH27 Living conditions | **0.773** | -0.007 | 0.093 | -0.157 |
| IPH30 Being taken seriously | **0.735** | -0.126 | 0.101 | 0.105 |
| IPH26 Feeling safe | **0.654** | 0.125 | 0.079 | 0.041 |
| IPH20 Being grateful | **0.653** | 0.137 | -0.015 | 0.196 |
| IPH16 Being high-spirited | **0.625** | 0.276 | -0.032 | 0.165 |
| IPH19 Accepting life | **0.618** | 0.158 | 0.021 | 0.150 |
| IPH14 Having control | **0.543** | 0.158 | 0.136 | 0.200 |
| IPH12 Accepting yourself | **0.543** | 0.248 | 0.106 | 0.067 |
| IPH34 Doing meaningful things | **0.531** | 0.120 | 0.081 | 0.210 |
| IPH4 Sleeping pattern | 0.332 | **0.695** | -0.084 | -0.194 |
| IPH2 Feeling fit | 0.057 | **0.687** | 0.141 | 0.161 |
| IPH6 Physical condition | -0.001 | **0.677** | 0.142 | 0.199 |
| IPH5 Eating pattern | 0.255 | **0.567** | 0.250 | -0.182 |
| IPH3 Having physical complaints or pain | -0.115 | **0.519** | 0.046 | 0.152 |
| IPH37 Knowing your limitations | 0.022 | 0.033 | **0.855** | -0.057 |
| IPH36 Looking after yourself | -0.088 | 0.117 | **0.791** | 0.030 |
| IPH40 Managing money | 0.106 | -0.032 | **0.775** | -0.082 |
| IPH39 Managing time | -0.022 | 0.062 | **0.772** | 0.057 |
| IPH42 Asking for help | 0.208 | -0.203 | **0.474** | 0.151 |
| IPH21 Continue learning | 0.090 | 0.020 | 0.044 | **0.758** |
| IPH17 Wanting to achieve ideals | 0.140 | 0.148 | -0.047 | **0.701** |
| IPH13 Being able to handle changes | 0.158 | -0.024 | 0.118 | **0.600** |

Extraction Method: Principal Component Analysis. Rotation Method: Oblimin with Kaiser Normalization. Pattern matrix, rotation converged in 7 iterations.

[1] During the process of item reduction the names of the factors were Contentment with life, Physical health and functioning, Self-management, and Personal development. Afterwards these were renamed by the research team into Contentment with life, Physical fitness, Daily life management, and Future perspective.
